# COVID-19 mortality in the Philippines: a province-level analysis, 2020-2023

**DOI:** 10.1101/2024.03.14.24304321

**Authors:** Jimuel Celeste, Jesus Emmanuel Sevilleja, Vena Pearl Bongolan, Roselle Leah Rivera, Salvador Eugenio Caoili, Romulo de Castro

## Abstract

**Objective:** To investigate the COVID-19 mortality in Philippine provinces from 2020 to 2023.

**Methods:** We calculated the crude (CMR) and age-standardised mortality rates (ASMR) of the Philippine provinces from the national COVID-19 surveillance data from January 18, 2020 to May 9, 2023. With Spearman’s method, we then performed a correlation analysis between the rates and four independent variables, namely, poverty incidence, population density, hospital beds per 100 000 population and proportion of elderly population (≥ 65).

**Results:** The province with the highest ASMR is Benguet with 207.83 deaths per 100 000 population, followed by Cagayan, Bataan, Nueva Vizcaya, Quirino, National Capital Region, Isabela, Cebu, Aurora and Davao del Sur. Meanwhile, the province with the lowest ASMR is Tawi-Tawi with 2.22 deaths per 100 000 population, followed by Sulu, Masbate, Misamis Occidental, Camarines Norte, Sarangani, Camiguin, Sultan Kudarat, Southern Leyte, Catanduanes and Batanes. Among the independent variables, poverty incidence and hospital beds per 100 000 population were found to independently predict CMR and ASMR.

**Discussion:** The results of this analysis provides a starting point for the investigation of COVID-19 mortality in Philippine provinces. The ranking of provinces by ASMR reveals the provinces with the highest and lowest rates, while the results of the correlation analysis, linked with findings from previous studies, explain the ranking, including surveillance issues that are related to health-care access and health-seeking behaviour. Our study paves the way for investigating factors that need to be considered and more closely investigated to get a clearer picture to inform decision-making, which affects millions of lives in communities across the Philippines’ provinces.

## INTRODUCTION

The World Health Organization (WHO) reports at least 6.9 million cumulative deaths from the coronavirus disease (COVID-19) globally.^1^ According to one report,^2^ this may even be an underestimation of the true death toll. With the lifting of the public health emergency of international concern status of COVID-19, it is time to look back, reflect and learn from our experience as the emergence of new viruses is expected.^3,4^ A better informed response to avoid preventable deaths in the future is possible.

In the Philippines, more than 66 000 deaths had been recorded as of May 2023.^5^ The Philippine Department of Health (DOH) defines a COVID-19 death as “a death resulting from a clinically compatible illness, in a probable or confirmed COVID-19 case, unless there is clear alternative cause of death that cannot be related to COVID-19 disease (e.g. trauma).”^6^ As further defined, a probable case satisfies both the clinical and the epidemiological criteria of COVID-19 infection or “death, not otherwise explained, in an adult with respiratory distress preceding death.”^6^ The clinical criteria pertains to the presence of influenza-like illness (cough and fever) and three of fatigue, headache, myalgia, sore throat, coryza, dyspnea, nausea, diarrhoea and anorexia. The epidemiological criteria is defined as “contact of a probable or confirmed case or linked to a COVID-19 cluster.”^6^ Meanwhile, a case is confirmed through RT-PCR test or rapid antigen test performed by a trained health professional if the former is not available. These definitions govern the COVID-19 mortality surveillance of the country.

This study provides a starting point for the investigation of COVID-19 mortality in Philippine provinces. As of this writing, no known studies have analysed the country’s province-level COVID-19 mortality data. We provide a ranking of provinces according to mortality rates and a list of variables that predicts the rates as well as issues surrounding the national COVID-19 surveillance data.

## METHODS

### Crude and age-standardised mortality rates

Province-level crude (CMR) and age-standardised mortality rates (ASMR)^7,8^ were computed for this analysis. Earlier global ranking studies argued that age-standardised rates should be used when ranking populations with varying age-structures to eliminate the age confounder.^7,8^ ASMR is suitable for the ranking of Philippine provinces which are known to have varying age distributions.^9^ ASMR allows the ranking of provinces while CMR provides an estimate of the true mortality rate.

Let ***i*** be the age group from ***0*** to ***n***, ***m*** be the number of deaths, ***p*** be the population, ***m / p*** be the age-specific mortality rate, and ***s*** be the standard population which in this study is the 2020 Philippine population.^9^ CMR and ASMR per 100 000 population are defined by the following equations:

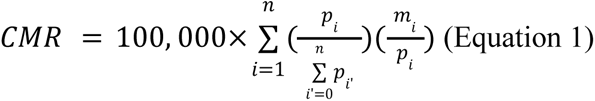

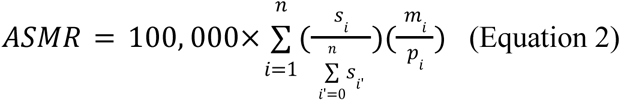

### COVID-19 data

We analyzed the national COVID-19 surveillance data of DOH.^5^ Records with missing age group, sex and province information were removed. From the 66 566 mortality records, 66 178 were retained after the filtering. Mortality by province and 5-year age group (0–4, …, ≥80) were then counted. 84 unique province values were found, 82 are provinces while 2 are cities (City of Isabela and Cotabato City). The dates of infection were then approximated from the date of disease onset, specimen collection, result release, report confirmation and death, whichever had a value, checked in the given order. The earliest case is dated January 18, 2020 while the latest is May 9, 2023. The mortality counts were used to calculate CMR and ASMR.

### Correlation Analysis

With CMR and ASMR calculated, we performed a correlation analysis to find variables that may predict the rates. We estimated Spearman’s rank correlation coefficients (**r**) between CMR, ASMR, difference between CMR and ASMR, and four independent variables: poverty incidence (2021),^10^ population density (2020),^11^ hospital beds per 100 000 population (2022)^9,12^ and proportion of elderly population (≥ 65).^9^ Poverty incidence is defined as “the proportion of Filipinos whose per capita income cannot sufficiently meet the individual basic food and non-food needs” which is “PhP 12,030 per month for a family of five” in 2021.^10^ Population density is “the number of persons per square kilometer of land.”^11^ Hospital beds per 100 000 population is the number of persons per hospital bed multiplied by 100 000. Proportion of elderly population is the number of elderly people (≥ 65) over the total population multiplied by 100. These independent variables represent the varying demographics, socioeconomic structures, and healthcare capacity of the provinces.

We used Spearman’s method to accommodate natural outliers.^13^ Moreover, we adopted the classification of Mukaka^13^ for coefficient interpretation: negligible correlation (0.00 to ±0.30), low (±0.30 to ±0.50), moderate (±0.50 to ±0.70), high (±0.70 to ±0.90) and very high (±0.90 to ±1.00). Finally, **P** values were calculated.

## RESULTS

### Crude and age-standardised mortality rates

The resulting CMR and ASMR are detailed in Table 1 and Figure 1. Ranking by ASMR shows that Benguet has the highest rate (207.83 deaths per 100 000 population) followed by the provinces of Cagayan, Bataan, Nueva Vizcaya, Quirino, National Capital Region (NCR), Isabela, Cebu, Aurora and Davao del Sur. Tawi-Tawi on the other hand has the lowest rate (2.22 deaths per 100 000 population) which is less than 1% of Benguet’s rate. It is followed in increasing order by Sulu, Masbate, Misamis Occidental, Camarines Norte, Sarangani, Camiguin, Sultan Kudarat, Southern Leyte, Catanduanes and Batanes. These provinces recorded the 10 highest and the 10 lowest ASMRs respectively.

**Figure 1.**
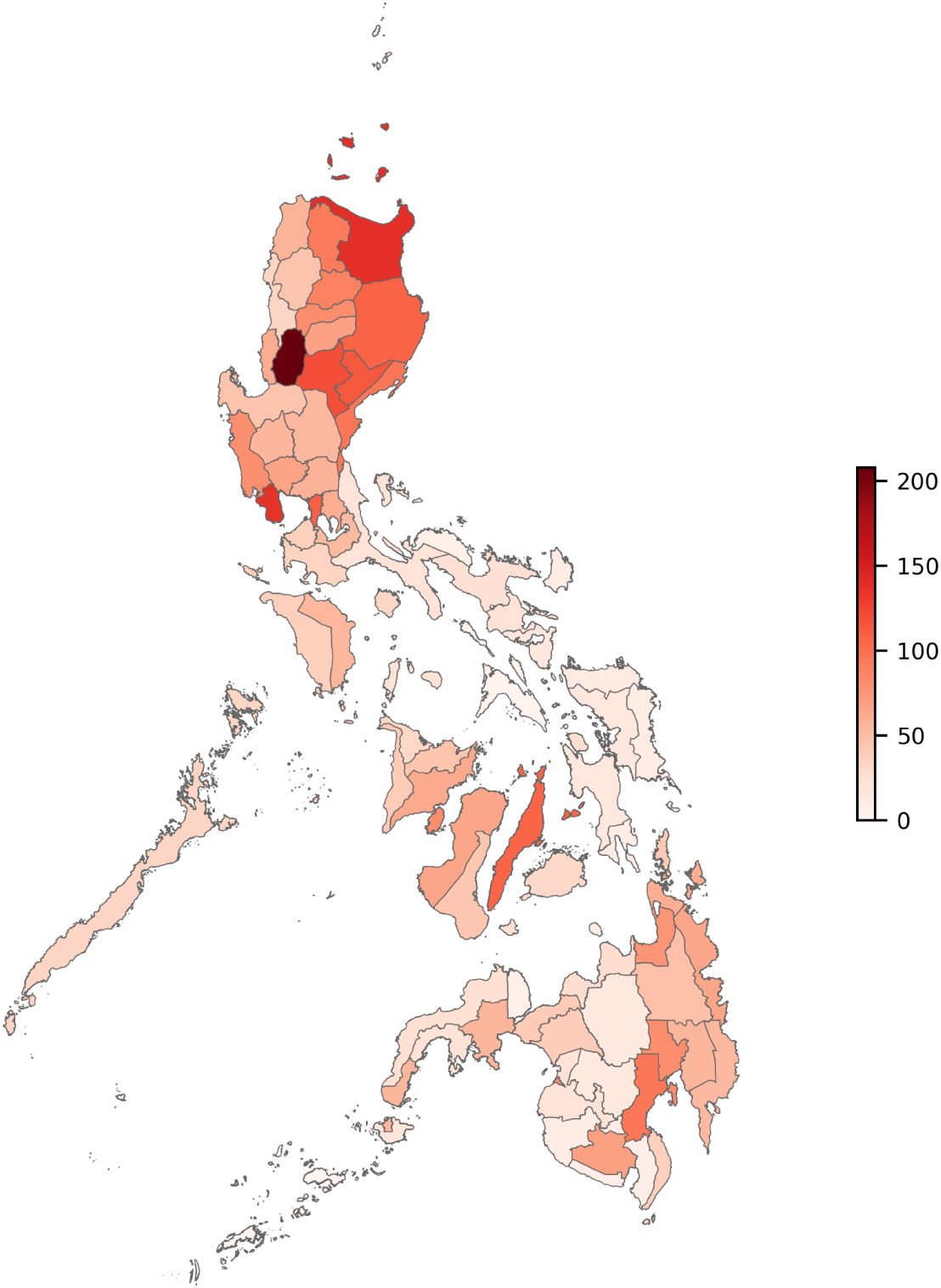
Heatmap of province-level age-standardized mortality rates per 100 000 population.

**Table 1.**
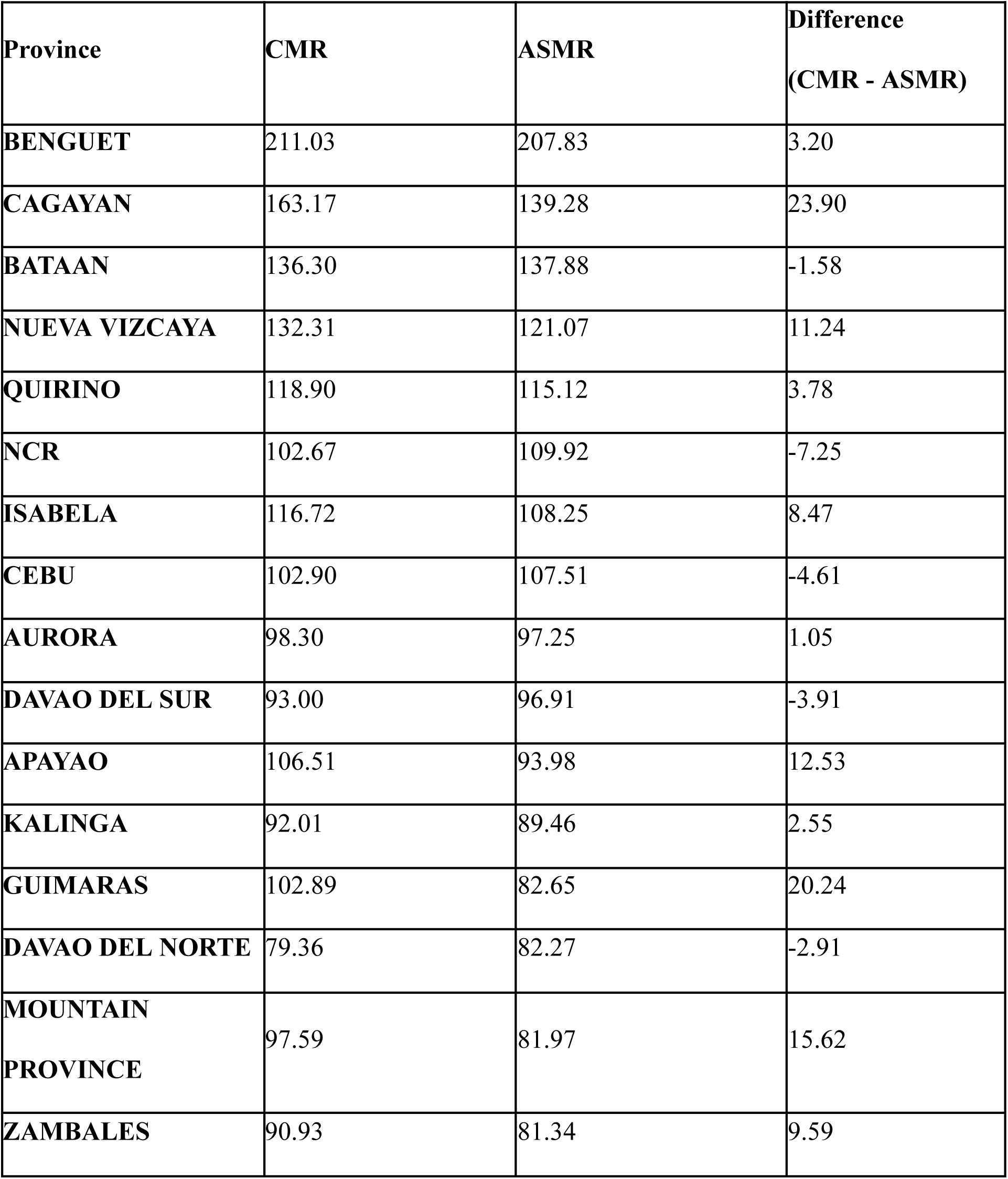
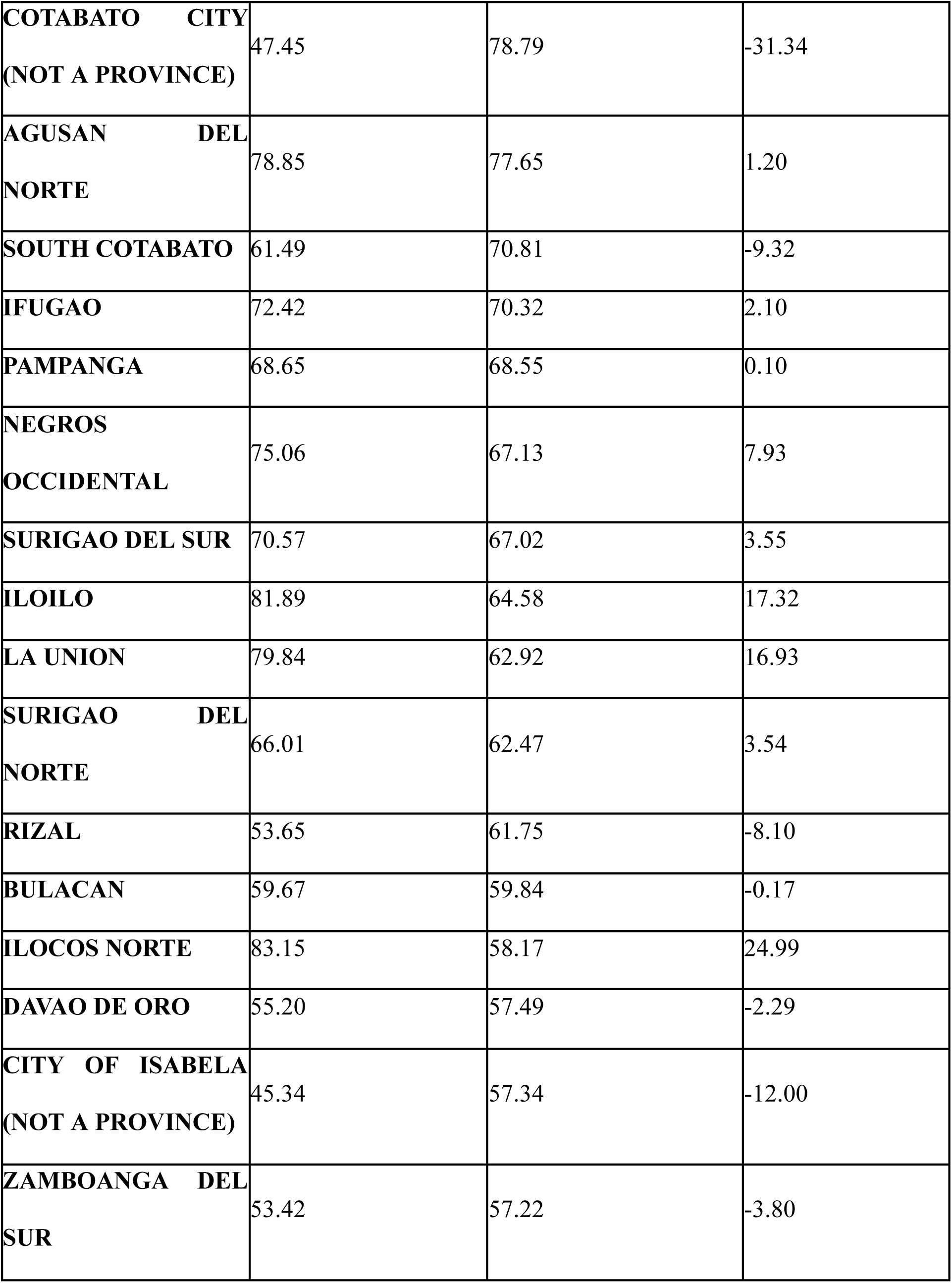
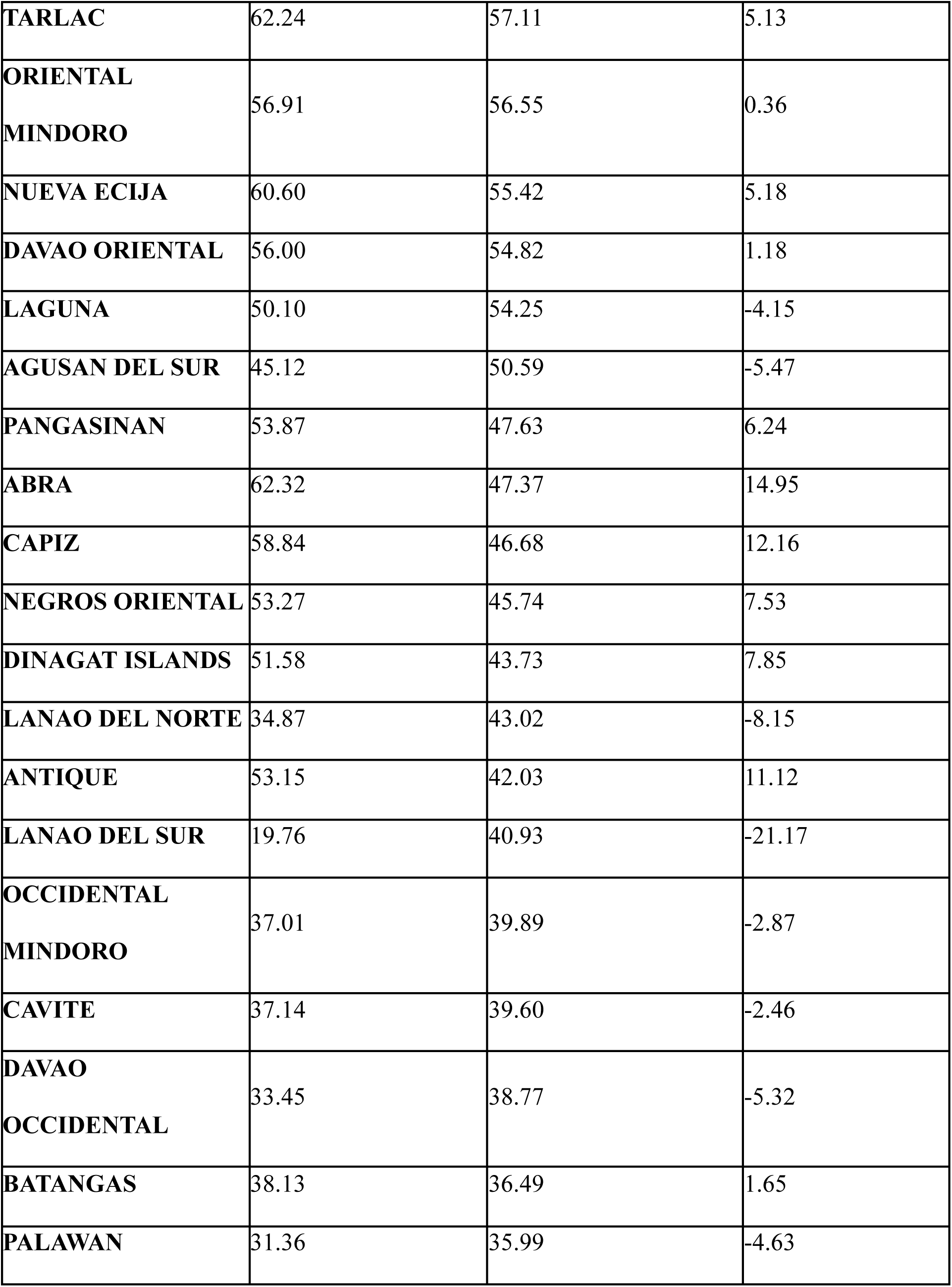
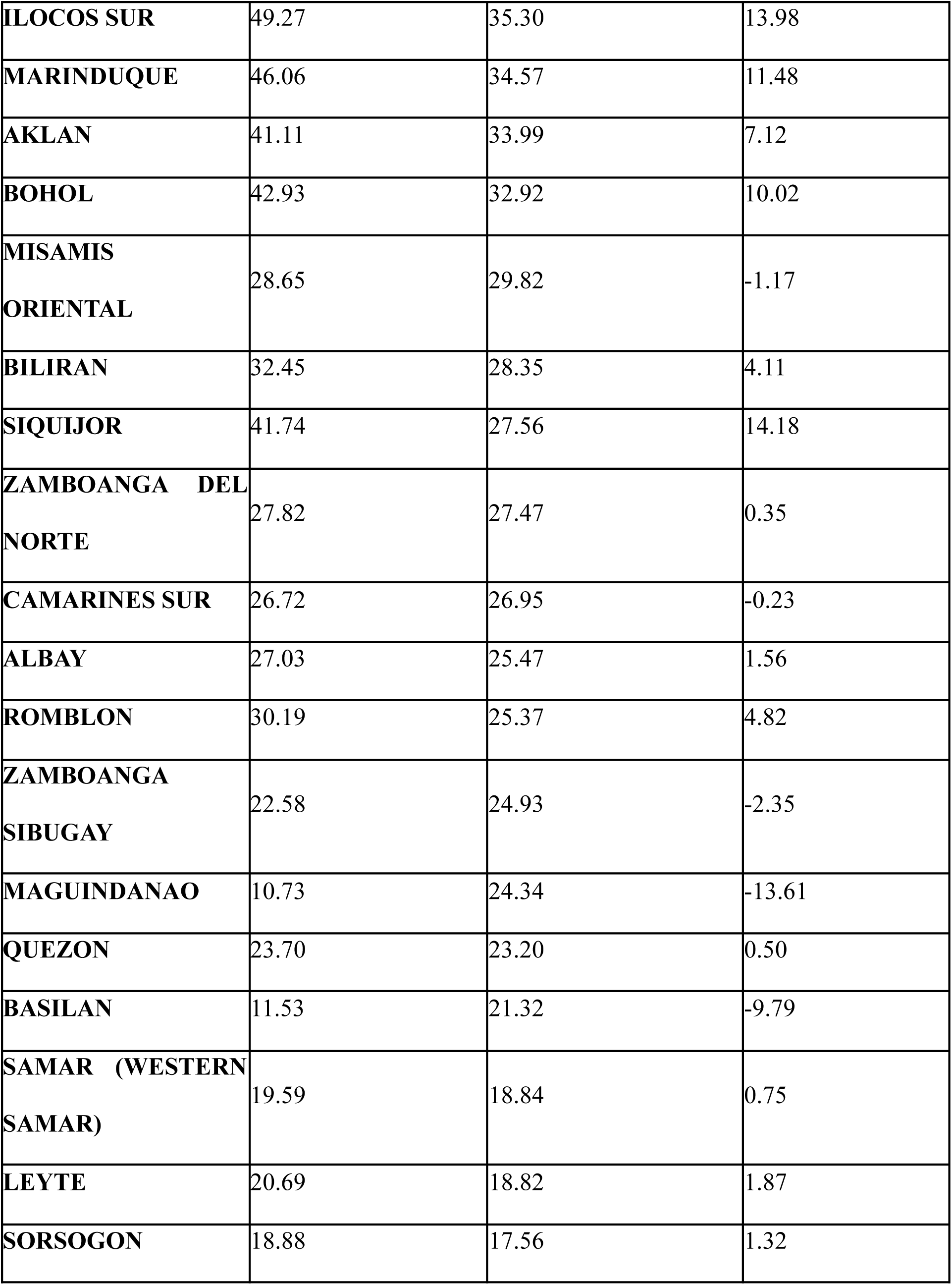
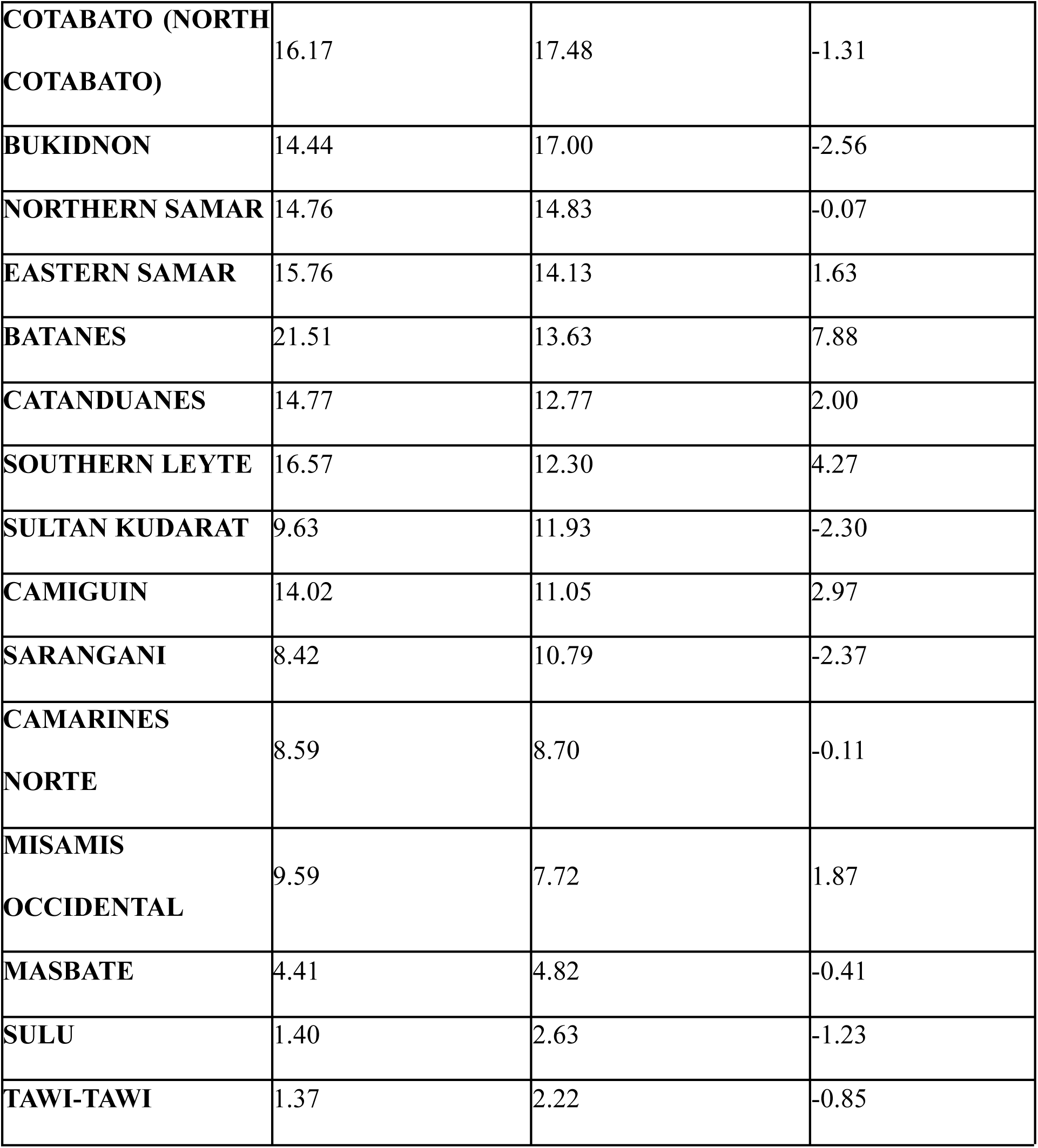
Province-level crude (CRM) and age-standardised mortality rates (ASMR) per 100 000 population, sorted by ASMR.

| Province | CMR | ASMR | Difference<br>(CMR - ASMR) |
| --- | --- | --- | --- |
| **BENGUET** | 211.03 | 207.83 | 3.20 |
| **CAGAYAN** | 163.17 | 139.28 | 23.90 |
| **BATAAN** | 136.30 | 137.88 | -1.58 |
| **NUEVA VIZCAYA** | 132.31 | 121.07 | 11.24 |
| **QUIRINO** | 118.90 | 115.12 | 3.78 |
| **NCR** | 102.67 | 109.92 | -7.25 |
| **ISABELA** | 116.72 | 108.25 | 8.47 |
| **CEBU** | 102.90 | 107.51 | -4.61 |
| **AURORA** | 98.30 | 97.25 | 1.05 |
| **DAVAO DEL SUR** | 93.00 | 96.91 | -3.91 |
| **APAYAO** | 106.51 | 93.98 | 12.53 |
| **KALINGA** | 92.01 | 89.46 | 2.55 |
| **GUIMARAS** | 102.89 | 82.65 | 20.24 |
| **DAVAO DEL NORTE** | 79.36 | 82.27 | -2.91 |
| **MOUNTAIN**<br>**PROVINCE** | 97.59 | 81.97 | 15.62 |
| **ZAMBALES** | 90.93 | 81.34 | 9.59 |
| **COTABATO CITY**<br>**(NOT A PROVINCE)** | 47.45 | 78.79 | -31.34 |
| **AGUSAN DEL**<br>**NORTE** | 78.85 | 77.65 | 1.20 |
| **SOUTH COTABATO** | 61.49 | 70.81 | -9.32 |
| **IFUGAO** | 72.42 | 70.32 | 2.10 |
| **PAMPANGA** | 68.65 | 68.55 | 0.10 |
| **NEGROS**<br>**OCCIDENTAL** | 75.06 | 67.13 | 7.93 |
| **SURIGAO DEL SUR** | 70.57 | 67.02 | 3.55 |
| **ILOILO** | 81.89 | 64.58 | 17.32 |
| **LA UNION** | 79.84 | 62.92 | 16.93 |
| **SURIGAO DEL**<br>**NORTE** | 66.01 | 62.47 | 3.54 |
| **RIZAL** | 53.65 | 61.75 | -8.10 |
| **BULACAN** | 59.67 | 59.84 | -0.17 |
| **ILOCOS NORTE** | 83.15 | 58.17 | 24.99 |
| **DAVAO DE ORO** | 55.20 | 57.49 | -2.29 |
| **CITY OF ISABELA**<br>**(NOT A PROVINCE)** | 45.34 | 57.34 | -12.00 |
| **ZAMBOANGA DEL**<br>**SUR** | 53.42 | 57.22 | -3.80 |
| **TARLAC** | 62.24 | 57.11 | 5.13 |
| **ORIENTAL**<br>**MINDORO** | 56.91 | 56.55 | 0.36 |
| **NUEVA ECIJA** | 60.60 | 55.42 | 5.18 |
| **DAVAO ORIENTAL** | 56.00 | 54.82 | 1.18 |
| **LAGUNA** | 50.10 | 54.25 | -4.15 |
| **AGUSAN DEL SUR** | 45.12 | 50.59 | -5.47 |
| **PANGASINAN** | 53.87 | 47.63 | 6.24 |
| **ABRA** | 62.32 | 47.37 | 14.95 |
| **CAPIZ** | 58.84 | 46.68 | 12.16 |
| **NEGROS ORIENTAL** | 53.27 | 45.74 | 7.53 |
| **DINAGAT ISLANDS** | 51.58 | 43.73 | 7.85 |
| **LANAO DEL NORTE** | 34.87 | 43.02 | -8.15 |
| **ANTIQUE** | 53.15 | 42.03 | 11.12 |
| **LANAO DEL SUR** | 19.76 | 40.93 | -21.17 |
| **OCCIDENTAL**<br>**MINDORO** | 37.01 | 39.89 | -2.87 |
| **CAVITE** | 37.14 | 39.60 | -2.46 |
| **DAVAO**<br>**OCCIDENTAL** | 33.45 | 38.77 | -5.32 |
| **BATANGAS** | 38.13 | 36.49 | 1.65 |
| **PALAWAN** | 31.36 | 35.99 | -4.63 |
| **ILOCOS SUR** | 49.27 | 35.30 | 13.98 |
| **MARINDUQUE** | 46.06 | 34.57 | 11.48 |
| **AKLAN** | 41.11 | 33.99 | 7.12 |
| **BOHOL** | 42.93 | 32.92 | 10.02 |
| **MISAMIS**<br>**ORIENTAL** | 28.65 | 29.82 | -1.17 |
| **BILIRAN** | 32.45 | 28.35 | 4.11 |
| **SIQUIJOR** | 41.74 | 27.56 | 14.18 |
| **ZAMBOANGA DEL**<br>**NORTE** | 27.82 | 27.47 | 0.35 |
| **CAMARINES SUR** | 26.72 | 26.95 | -0.23 |
| **ALBAY** | 27.03 | 25.47 | 1.56 |
| **ROMBLON** | 30.19 | 25.37 | 4.82 |
| **ZAMBOANGA**<br>**SIBUGAY** | 22.58 | 24.93 | -2.35 |
| **MAGUINDANAO** | 10.73 | 24.34 | -13.61 |
| **QUEZON** | 23.70 | 23.20 | 0.50 |
| **BASILAN** | 11.53 | 21.32 | -9.79 |
| **SAMAR (WESTERN**<br>**SAMAR)** | 19.59 | 18.84 | 0.75 |
| **LEYTE** | 20.69 | 18.82 | 1.87 |
| **SORSOGON** | 18.88 | 17.56 | 1.32 |
| **COTABATO (NORTH COTABATO)** | 16.17 | 17.48 | -1.31 |
| **BUKIDNON** | 14.44 | 17.00 | -2.56 |
| **NORTHERN SAMAR** | 14.76 | 14.83 | -0.07 |
| **EASTERN SAMAR** | 15.76 | 14.13 | 1.63 |
| **BATANES** | 21.51 | 13.63 | 7.88 |
| **CATANDUANES** | 14.77 | 12.77 | 2.00 |
| **SOUTHERN LEYTE** | 16.57 | 12.30 | 4.27 |
| **SULTAN KUDARAT** | 9.63 | 11.93 | -2.30 |
| **CAMIGUIN** | 14.02 | 11.05 | 2.97 |
| **SARANGANI** | 8.42 | 10.79 | -2.37 |
| **CAMARINES NORTE** | 8.59 | 8.70 | -0.11 |
| **MISAMIS OCCIDENTAL** | 9.59 | 7.72 | 1.87 |
| **MASBATE** | 4.41 | 4.82 | -0.41 |
| **SULU** | 1.40 | 2.63 | -1.23 |
| **TAWI-TAWI** | 1.37 | 2.22 | -0.85 |

Comparing CMR and ASMR (Tables 1 and 2), CMR has a higher median and wider dispersion (48.36 deaths per 100 000 (IQR 22.32 - 73.08)) than ASMR (44.73 deaths per 100 000 (IQR 24.78 - 67.48)). The median difference between the two variables is 1.26 deaths per 100 000 (IQR -2.35 - 7.22). Ilocos has the highest positive difference while Cotabato City has the highest negative difference: 24.99 and -31.34 deaths per 100 000 population respectively. In the context of ranking, these differences are not negligible, demonstrating the utility and importance of age-standardisation when comparing populations with varying age structures.

**Table 2.**
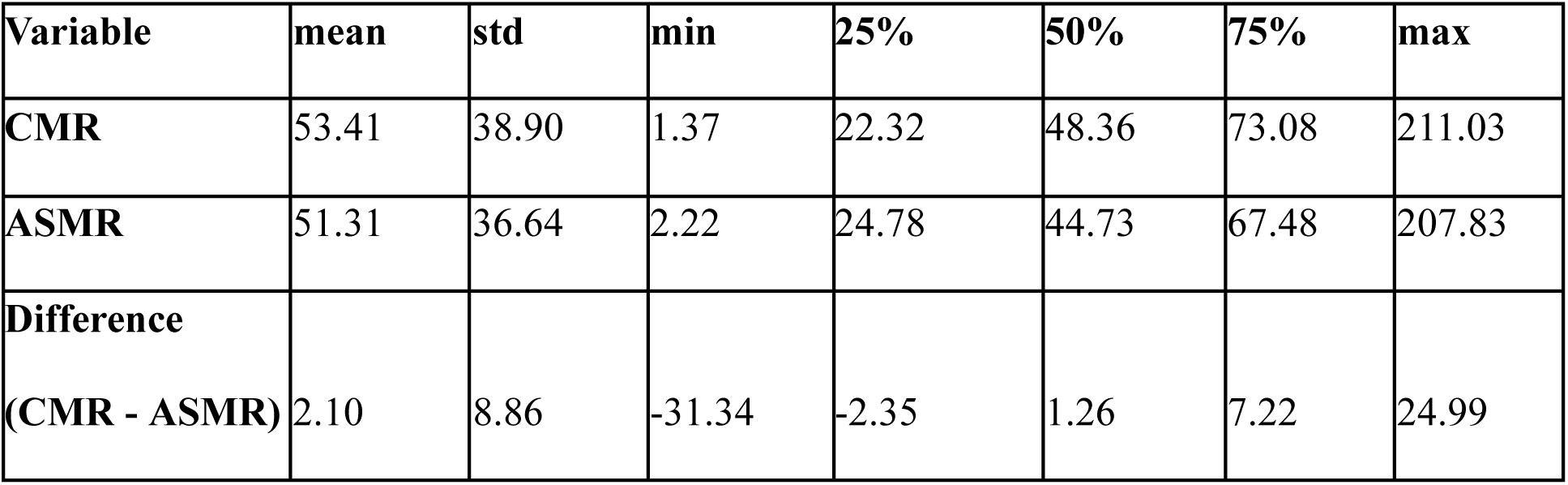
Summary of province-level CMR and ASMR per 100 00 population.

The age-specific mortality rates per 100 000 are detailed in Figure 2 and Table 3. The distribution follows a check-shaped pattern, highlighting the steep increase in mortality rate as age increases and a noticeable uptick in the infant age group (0-4). The infant age group has a higher median (4.79 deaths per 100 000 (IQR 2.15 - 9.61)) compared to the adjacent age groups 5-9, 10-14 and 15-19. In fact, age group 5-9 recorded the lowest median value at 0.53 deaths per 100 000 (IQR 0 - 1.95). The highest value, in contrast, is recorded in the oldest age group (≥80): 640.90 deaths per 100 000 (IQR 347.27 - 1046.65). To put into perspective, the highest value is 1209.25-fold higher than the lowest value. This difference shows the stark contrast in COVID-19 mortality risk between younger people and the elderly.

**Figure 2.**
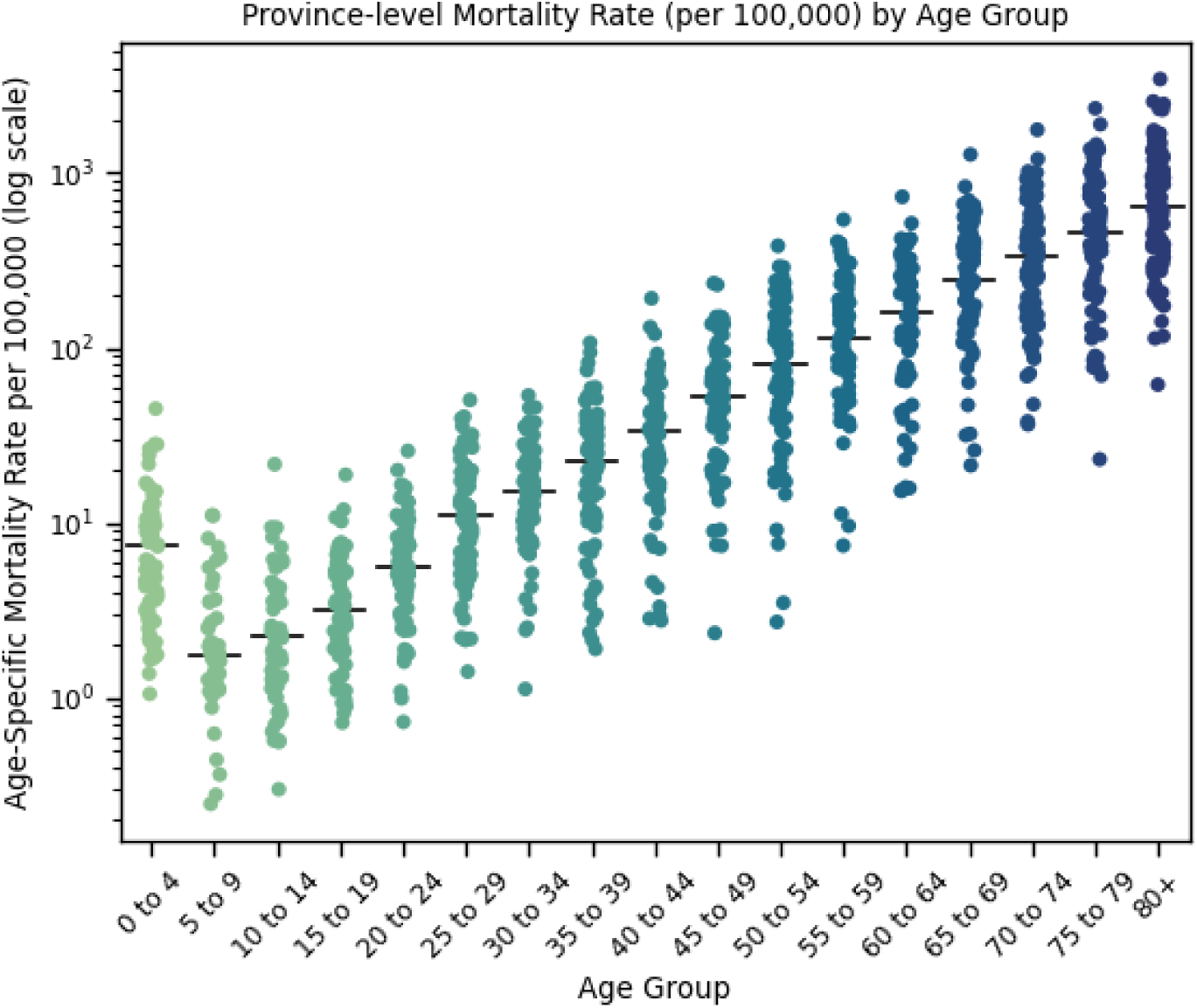
Strip plot (with median markers) of province-level age-specific mortality rates per 100 000 population.

**Table 3.** Summary of province-level age-specific mortality rates per 100 000 population.

| **Age Group** | **mean** | **std** | **min** | **25%** | **50%** | **75%** | **max** |
| --- | --- | --- | --- | --- | --- | --- | --- |
| **0 to 4** | 7.22 | 7.81 | 0.00 | 2.15 | **4.79** | 9.61 | 45.07 |
| **5 to 9** | 1.51 | 2.34 | 0.00 | 0.00 | **0.53** | 1.95 | 11.06 |
| **10 to 14** | 2.14 | 3.10 | 0.00 | 0.00 | **1.31** | 2.42 | 21.65 |
| **15 to 19** | 2.91 | 3.36 | 0.00 | 0.00 | **1.95** | 4.47 | 18.85 |
| **20 to 24** | 6.08 | 4.94 | 0.00 | 2.49 | **5.12** | 8.42 | 25.79 |
| **25 to 29** | 11.74 | 10.71 | 0.00 | 4.77 | **8.67** | 16.50 | 50.44 |
| **30 to 34** | 15.78 | 11.82 | 0.00 | 7.84 | **14.35** | 21.51 | 53.59 |
| **35 to 39** | 24.07 | 21.55 | 0.00 | 7.40 | **20.70** | 33.98 | 107.36 |
| **40 to 44** | 35.44 | 31.63 | 0.00 | 15.24 | **29.84** | 46.04 | 192.31 |
| **45 to 49** | 58.63 | 48.08 | 0.00 | 23.96 | **46.23** | 69.63 | 234.53 |
| **50 to 54** | 92.27 | 75.86 | 0.00 | 39.89 | **64.83** | 120.15 | 384.16 |
| **55 to 59** | 145.48 | 104.88 | 0.00 | 76.67 | **124.86** | 183.13 | 542.28 |
| **60 to 64** | 195.86 | 136.76 | 0.00 | 92.27 | **187.60** | 271.82 | 732.91 |
| **65 to 69** | 305.77 | 206.24 | 0.00 | 157.96 | **284.43** | 391.84 | 1279.52 |
| **70 to 74** | 435.95 | 311.59 | 36.57 | 194.27 | **364.22** | 593.63 | 1769.76 |
| **75 to 79** | 545.14 | 442.88 | 0.00 | 211.89 | **472.67** | 736.99 | 2350.30 |
| **80+** | 792.22 | 640.48 | 0.00 | 347.27 | **640.90** | 1046.65 | 3453.26 |

### Correlation analysis

From the correlation analysis (Tables 4 and 5 and Figure 3), two variables independently predict COVID-19 mortality rates: poverty incidence and hospital beds per 100 000 population. Poverty incidence has a moderate negative correlation with CMR and low negative correlation with ASMR, while hospital beds per 100 000 population has a low positive correlation with both CMR and ASMR. Meanwhile, there is no correlation found between the mortality rates and the two other independent variables: population density and proportion of elderly population (≥65). Note however that the correlation coefficient between the proportion of elderly population and CMR (***r*** = 0.21) is higher than that of ASMR which is almost zero (***r*** = -0.01) due to age-standardisation.

**Figure 3.**
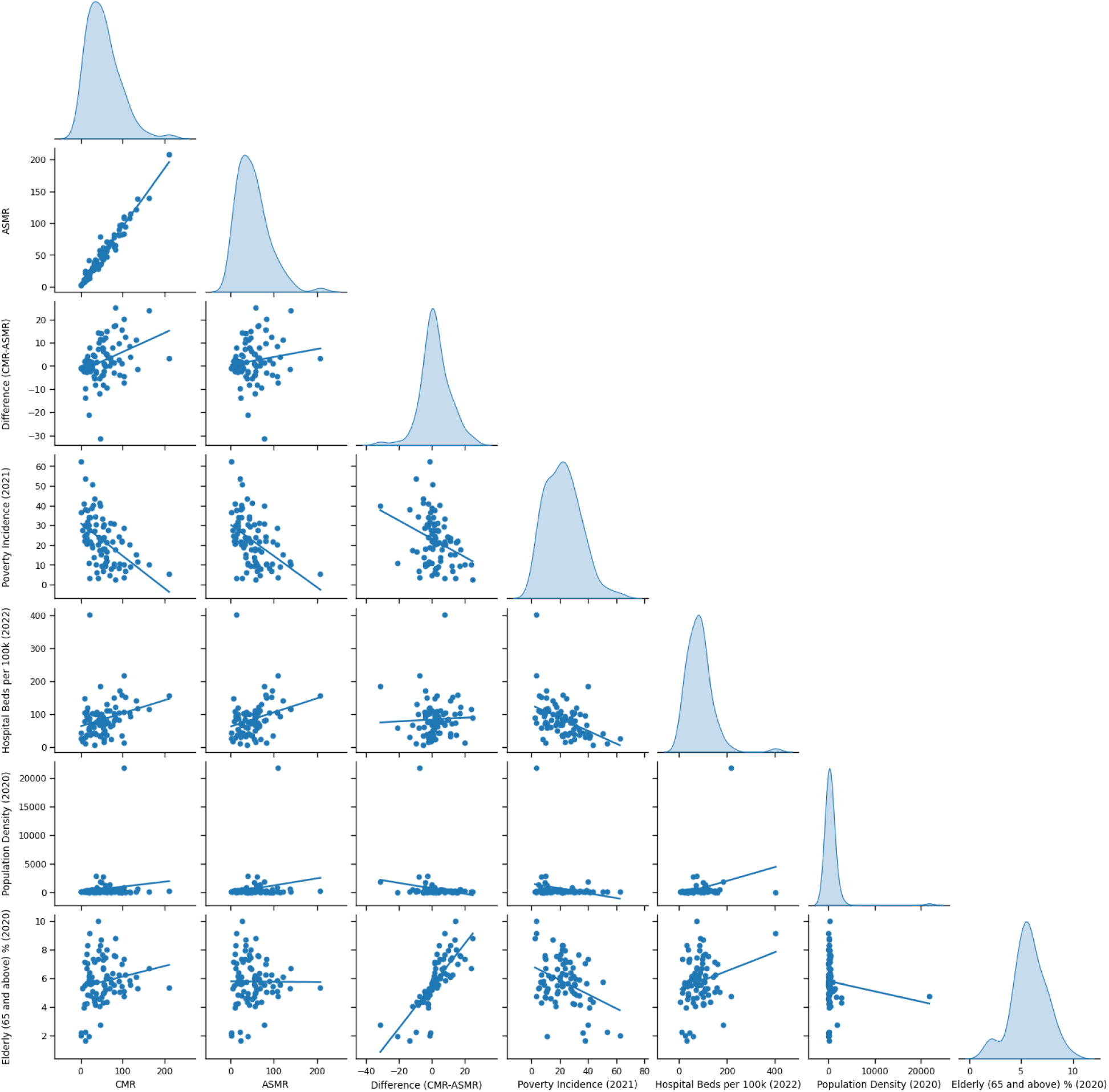
Scatter plot with regression line and kernel density estimate diagonal plot of CMR, ASMR, the difference between CMR and ASMR, and the four independent variables: poverty incidence, population density, hospital beds per 100 000 population, and proportion of elderly population (≥65)

**Table 4.** Spearman’s rank correlation coefficient and P-value of CMR, ASMR, the difference between CMR and ASMR, and the four independent variables: poverty incidence, population density, hospital beds per 100 000 population, and proportion of elderly population (≥65)

| Dependent Variables | Poverty Incidence (2021) | | Population Density (2020) | | Hospital Beds per 100 000 Population (2022) | | Proportion of Elderly Population ($\geq 65$) (2020) | |
| --- | --- | --- | --- | --- | --- | --- | --- | --- |
|  | r | P | r | P | r | P | r | P |
| **CMR** | -0.55 | < 0.001 | 0.10 | 0.204 | 0.35 | < 0.05 | 0.21 | 0.109 |
| **ASMR** | -0.49 | < 0.001 | 0.11 | 0.081 | 0.33 | <0.05 | -0.01 | 0.962 |
| **Difference (CMR - ASMR)** | -0.30 | < 0.05 | -0.12 | 0.109 | 0.13 | 0.686 | 0.90 | < 0.001 |

**Table 5.**
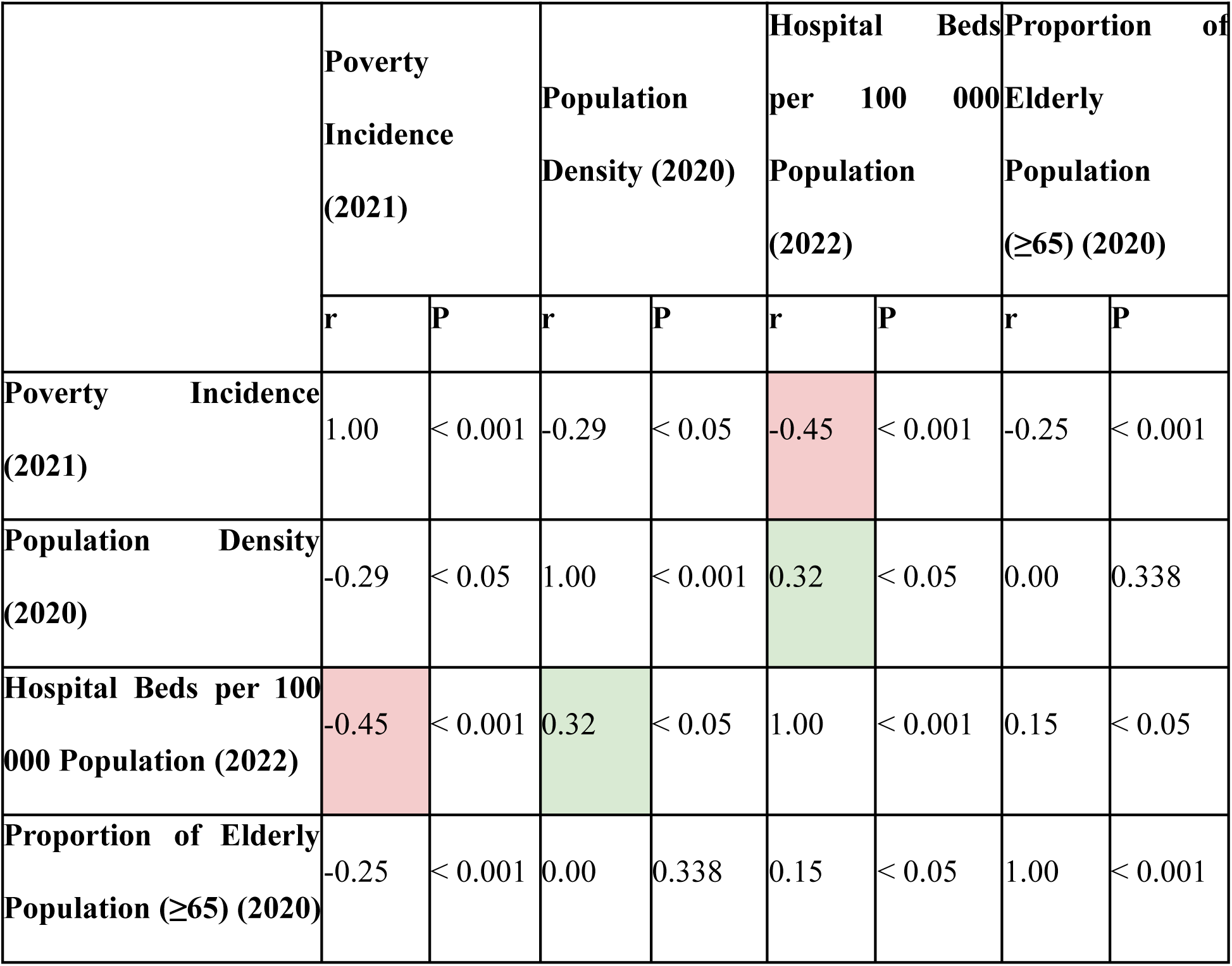
Spearman’s rank correlation coefficient and P-value of the four independent variables: poverty incidence, population density, hospital beds per 100 000 population, and proportion of elderly population (≥65)

| | Poverty Incidence (2021) | | Population Density (2020) | | Hospital Beds per 100 000 Population (2022) | | Proportion of Elderly Population ($\geq 65$) (2020) | |
| --- | --- | --- | --- | --- | --- | --- | --- | --- |
|  | r | P | r | P | r | P | r | P |
| **Poverty Incidence (2021)** | 1.00 | < 0.001 | -0.29 | < 0.05 | -0.45 | < 0.001 | -0.25 | < 0.001 |
| **Population Density (2020)** | -0.29 | < 0.05 | 1.00 | < 0.001 | 0.32 | < 0.05 | 0.00 | 0.338 |
| **Hospital Beds per 100 000 Population (2022)** | -0.45 | < 0.001 | 0.32 | < 0.05 | 1.00 | < 0.001 | 0.15 | < 0.05 |
| **Proportion of Elderly Population ($\geq 65$) (2020)** | -0.25 | < 0.001 | 0.00 | 0.338 | 0.15 | < 0.05 | 1.00 | < 0.001 |

Associations between the independent variables were also found. Hospital beds per 100 000 population has a low negative correlation with poverty incidence and low positive correlation with population density.

## DISCUSSION

### Ranking of provinces by age-standardised mortality rates

Our analysis provides a ranking of provinces by ASMR and a list of variables that predict the rates. The provinces with the highest rates are Benguet, Cagayan, Bataan, Nueva Vizcaya, Quirino, NCR, Isabela, Cebu, Aurora and Davao del Sur. Eight of these provinces are in Luzon while the other two, Cebu and Davao del Sur, are in Visayas and Mindanao respectively. The provinces of Benguet, NCR, Cebu and Davao del Sur are also known to house densely populated cities: Baguio City in Benguet; 16 cities in NCR; the cities of Cebu, Lapu-Lapu and Mandaue in Cebu; and Davao City in Davao del Sur. Meanwhile, the provinces with the lowest rates are Tawi-Tawi, Sulu, Masbate, Misamis Occidental, Camarines Norte, Sarangani, Camiguin, Sultan Kudarat, Southern Leyte, Catanduanes and Batanes. The provinces of Tawi-Tawi, Sulu, Catanduanes and Batanes are composed of smaller islands and are relatively remote compared to the rest of the provinces.

With these enumerated characteristics, we can speculate that population density and geographical connectedness may have contributed in COVID-19 mortality by means of faster transmission and hence higher number of infections. However, we found negligible correlation between population density and CMR or ASMR. The contribution of these factors to mortality rate cannot be ruled out but the data does not provide evidence to support it. Instead, we found correlations with poverty incidence and hospital beds per 100 000 population.

### Poverty incidence and hospital beds per 100 000 population are predictors

CMR and ASMR tends to be higher in provinces with lower poverty incidence. This relationship is counter-intuitive. COVID-19 disproportionately affects low-income populations due to overcrowding, working conditions that do not allow work-from-home setups (work mobility), financial uncertainty, disproportionate health-care access and more advanced comorbidities which are undiagnosed or untreated.^14^ Evidence from Sweden supports this argument by showing that low income, low education level and being born in a low- or middle-income country predicts higher risk of death from COVID-19.^15^ Hence, the counter-intuitive relationship might not be suggesting that provinces with high poverty incidence have a better chance of surviving COVID-19. We hypothesize that it is a surveillance issue.

CMR and ASMR tends to be higher in provinces with higher number of hospital beds per 100 000 population. The definition of a probable and a confirmed COVID-19 case in the Philippines requires clinical determination and so deaths that happened in hospitals or healthcare facilities with testing capacity might have been recorded better than cases outside healthcare facilities. Moreover, we also found that hospital beds per 100 000 tend to be lower in provinces with higher poverty incidence. Taken together, these relationships may be suggesting that CMR and ASMR tends to be lower in provinces with higher poverty incidence because hospital beds tend to be lower in these provinces, which may have left many COVID-19 deaths unrecorded.

### The streetlight effect

The phenomenon at which cases are higher in populations with better surveillance is coined as the “streetlight effect.”^16^ In the early stage of the pandemic in 2020, the streetlight effect was observed in the regional COVID-19 data of Western Visayas, Philippines as reported in a preprint by Zamora et al. (Zamora PRF, Rico JA, Bolinas DK, Sevilleja JE, de Castro R. 2022. Early COVID-19 pandemic response in Western Visayas, Philippines. medRxiv. doi:10.1101/2022.07.21.22277909). They analysed the testing, infection and contact tracing data in Western Visayas and have found that the highest number of cases is recorded in the City of Iloilo which houses the only regional COVID-19 testing facility in Western Visayas at that time. We speculate that the national COVID-19 surveillance data also exhibits the streetlight effect.

Take the case of Benguet. Benguet has the highest computed ASMR with 207.83 deaths per 100 000 population. To put into perspective, the province next to it, Cagayan, has an ASMR value of 139.28 deaths per 100 000 population. Benguet is an outlier and it may be linked to the province’s extensive testing and contact tracing system. In 2020, Baguio City and the province of Benguet overall were commended by WHO for their exceptional COVID-19 pandemic response in terms of contact tracing and related control measures.^17^ The contact tracing system of the province allowed it to trace and record infections and mortalities perhaps better than other provinces that do not have a similarly extensive system in place.

However, this is not to say that testing and contact tracing are the only factors that may have contributed to the high mortality rate of Benguet. The relatively low temperature of Benguet may also be linked to its higher ASMR. Baguio City has a mean annual temperature of 18 °C,^18^ while all other weather stations in the country have a mean annual temperature of 26 °C,^18^ making it the coldest city in the Philippines on average. Ecological studies^19–21^ found associations between temperature, humidity and COVID-19 mortality rate: colder and humid geographies record worse cases and higher mortalities. Furthermore, Benguet houses Baguio City which is densely populated. Extensive testing and contact tracing, population density, and climate are among the factors that might have contributed to the high mortality rate of the province.

In addition to the availability and extent of surveillance efforts, health-seeking behaviour also contributes to the numbers. It was found in a repeated cross-sectional survey in low-income households in the Philippines that the “intention to seek care from public hospitals for COVID-19 symptoms dropped from 43.6% to 28.4%, while reports of self-treatment using stored medicines or antibiotics increased.”^22^ Low-income households already have disproportionate access to health care and still have been found to have low health-seeking behaviour during the pandemic.

### Check-shaped age-specific mortality rates

We found negligible correlation between the proportion of elderly population and CMR and an almost zero correlation with ASMR which is expected due to age-standardisation. However, our analysis of the age-specific mortality rates shows a check-shaped pattern which highlights the increasing COVID-19 mortality risk as age increases. The more interesting detail is the uptick observed in the infant age group (0-4). This pattern was previously reported in an analysis of age-specific COVID-19 mortality data of US, UK and Spain.^23^ The researchers reported a “U-shaped” pattern in the infant and children’s age group: “Although it is commonly believed that children are completely spared of COVID-19, our findings suggest that newborns and children during their first year of life exhibit a slightly elevated COVID-19 risk.”^23^ The results of our analysis add the Philippines to the list of countries where such a pattern is observed.

### Limitations

Our study is without its limitations. First, correlation analysis ignores the local dynamics within provinces. A closer investigation of provinces is necessary to identify factors and interventions that worked best or poorly within the provinces. Second, several important variables are not included in our study, namely, tests performed by province, government policies such as travel restrictions and lockdowns, climate variables and comorbidities within provinces. Third, the temporal dimension of the data is not considered in the analysis. Future studies may perform correlation analysis on several periods during the pandemic. A variable may predict mortality rates at one point and not in another due to changing context over time. Finally, the ranking of provinces may be influenced by the streetlight effect as we hypothesised. The ranking may be improved with the re-estimation of COVID-19 deaths through excess deaths.^24^ Despite these limitations, our study paves the way for investigating factors that need to be considered and more closely investigated to get a clearer picture to inform decision-making, which affects millions of lives in communities across the Philippines’ provinces.

### Summary

Our study provides the first province-level analysis of COVID-19 mortality in Philippine provinces. We computed the crude and age-standardised mortality rates of the Philippine provinces. We then ranked the provinces with the age-standardised rates. A correlation analysis was then performed to find variables that predict the rates. We found that poverty incidence and hospital beds per 100 000 population independently predict mortality rates. Linking our findings with knowledge from previous studies, we also fleshed out COVID-19 surveillance issues such as the streetlight effect and how it is related with health-care access and health-seeking behaviour, which are deeper than testing and contact tracing alone. Our study takes a thorough look at the need to unpack specific factors and their relationships with mortality rates in the provinces that have not been explored in the past.

## Data Availability

The publicly available national COVID-19 surveillance data used for this study was last accessed on December 13, 2023 from https://doh.gov.ph/covid19tracker. Upon checking on March 14, 2024, the link is no longer accessible.

## ACKNOWLEDGMENTS

The authors thank the Office of the Vice Chancellor for Research and Development, University of the Philippines Diliman through their SOS grants for their support of our COVID-19 studies.

## ETHICS STATEMENT

Ethical review and approval was not required for the study. The data used for analysis were downloaded from publicly available sources.

